# Oxidative Stress, Neuroinflammation, and Neuronal Vulnerability Begin in Midlife: a 7 Tesla Magnetic Resonance Spectroscopy Healthy Adult Lifespan Study

**DOI:** 10.1101/2025.10.19.25338308

**Authors:** Flavie E. Detcheverry, Sneha Senthil, Rozalia Arnaoutelis, Samson Antel, Haz-Edine Assemlal, Zahra Karimaghaloo, Douglas L. Arnold, Jamie Near, Sridar Narayanan, AmanPreet Badhwar

## Abstract

**INTRODUCTION:** While changes associated with age-related diseases, like oxidative stress, begin in midlife, most aging studies focused on older individuals. Our study assessed *in vivo* brain metabolites in healthy adults, including the understudied middle-age group.

**METHODS:** 7 tesla magnetic resonance spectroscopy data were acquired from 95 healthy adults (48 women) aged 20-79 years. Eight metabolites were measured in posterior cingulate cortex (PCC) and centrum semiovale white matter (CSWM).

**RESULTS:** With increasing age, we found (a) lower glutathione and glutamate, and higher *myo-*inositol in PCC, and (b) lower *N*-acetylaspartate and glutamate, and higher *myo*-inositol, total creatine, and *N*-acetylaspartyl-glutamate in CSWM. Notably, most changes started in midlife and were driven by age-related changes in women.

**DISCUSSION:** Overall, we found evidence that oxidative stress, neuroinflammation, and neuronal vulnerability begin in midlife in healthy adults. Targeting these processes in midlife may slow brain aging and reduce age-related neurodegenerative diseases risk, including Alzheimer’s disease.

**GRAPHICAL ABSTRACT:** 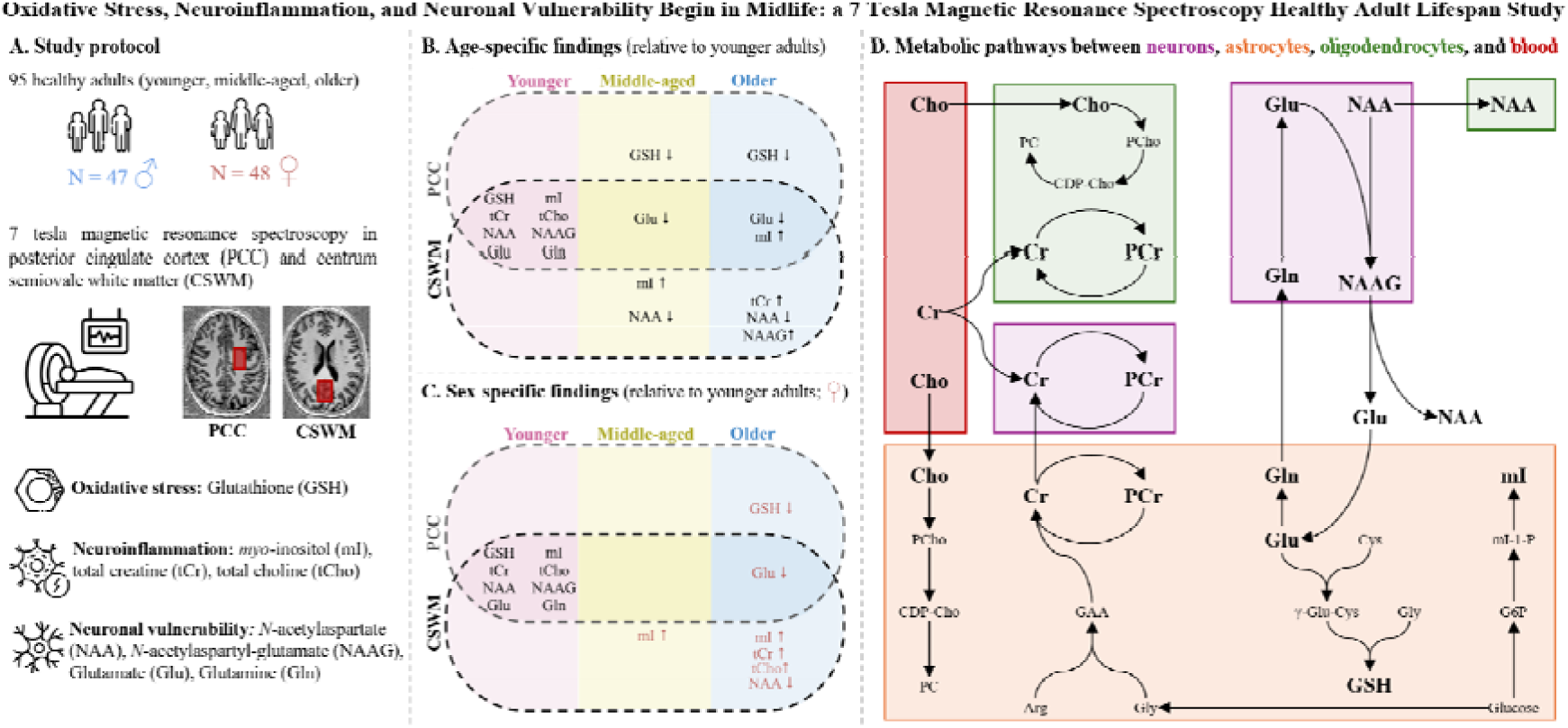

## 1. INTRODUCTION

Higher life expectancy is accompanied by higher prevalence of age-related neurodegenerative diseases (NDDs), with Alzheimer’s disease (AD) being the most prevalent [1]. Evidence suggests that NDD-associated structural, functional, and molecular brain changes begin in midlife (40+ years) [2,3], and might be accelerated by menopause in women [2]. Therefore, the transition from middle- to older-age is crucial for future health and might offer a window for cognitive decline prevention. Notably, the 2024 *Lancet’s* Commission on Dementia reported that addressing 14 identified risk factors could prevent 45% of dementia cases, with ten, including metabolic health, occurring in midlife and potentially reducing dementia risk by 30% [4]. Despite this, most aging- and NDD-studies focused on older individuals (60+ years).

Oxidative stress (OS) has been hypothesized as a key contributor to age-related decline [5,6]. OS results from an imbalance between the production and neutralization of reactive oxygen species (ROS), like hydrogen peroxide [7]. This imbalance is amplified in midlife as ROS accumulate, gradually weakening antioxidant defenses and disrupting homeostasis [8], with menopause further increasing oxidative stress in women [9]. Being the most energetically demanding organ [10], the brain, and especially hub regions like the posterior cingulate cortex (PCC) [11,12], are particularly vulnerable to OS. Antioxidants, with the most prevalent being the metabolite glutathione (GSH), can help neutralize ROS and protect against oxidative damage [6]. GSH and other metabolites can be assessed in the brain *in vivo* using magnetic resonance spectroscopy (MRS). GSH levels were found to (a) decrease with age in posterior regions in older relative to younger adults and in younger-to-middle-aged adults as one group, reflecting lower antioxidant capacity, and (b) increase in frontal regions in older relative to younger adults, suggesting a compensatory mechanism against increased OS [7]. GSH differences with middle-aged healthy adults as a distinct group, however, have not been investigated with MRS.

In addition to GSH reflecting antioxidant capacity, additional metabolites can be assessed *in vivo* using MRS, reflecting neuroinflammatory and neurodegenerative processes. MRS-detected metabolites most studied include *N*-acetylaspartate (NAA; neuroaxonal marker); *N*-acetylaspartyl-glutamate (NAAG; neuromodulator); glutamate (Glu; main excitatory neurotransmitter); choline (Cho; membrane turnover marker); *myo*-inositol (mI; gliosis marker); and creatine (Cr; energy substrate) [13]. Although decreased NAA and Glu, and increased Cho, mI, and Cr levels have commonly been reported with increasing age, there lacks consensus [14–16], with variable ages and sample sizes across studies being a likely contributor. Specifically, studies investigating middle-aged healthy adults are lacking, an age group where changes enhancing risks of developing an NDD later in life are thought to appear [17,18].

Other reasons might explain the lack of consensus and increase the variability of MRS findings. Notably, the variety of MRS parameters and quantification techniques (e.g., ratios to Cr) can influence metabolite detection and complicate results interpretation. For example, using 1.5 tesla (T), McIntyre et al. reported higher total NAA and total Cr levels in centrum semiovale white matter (CSWM) using short echo time (TE), and no significance using long TE [19]. There is also a scarcity of 7T MRS studies, a magnet strength providing better signal-to-noise ratio, improved spectral resolution [20,21], and excellent reproducibility of low-concentration metabolites measurements like GSH [7,21]. Finally, most age-related metabolite findings are region-specific [7,22], which might be due to the underlying tissue composition, as some metabolites are preferentially located in certain cell types (e.g., GSH in glia).

Therefore, we assessed brain metabolite level variations in healthy adults, including midlife, using 7T MRS in PCC and CSWM, two areas showing altered microstructure, metabolism, and connectivity in aging and NDDs (including AD) [11,12,23,24]. Given the increased sensitivity and specificity provided by ultra-high field (UHF) strength, ultra-short TE, and long repetition time (TR) [25], we were able to reliably measure levels of eight metabolites, including three low-concentration metabolites (GSH, NAAG, glutamine [Gln]). Specifically, we assessed the associations between metabolite levels and age, sex, and functional tests.

## 2. METHODS

### 2.1 Participants

Ninety-five healthy adults (48 women) belonging to three age groups were included: younger, 20-39 years, N = 32; middle-aged, 40-59 years, N = 32; older, 60-79 years, N = 31. Participants were recruited from the Montreal community using the word-of-mouth method, as well as from the *Banque de Participants* (https://criugm.qc.ca/la-recherche-jy-participe/conferences-de-la-banque-de-participants/) established at the *Centre de Recherche de l’Institut Universitaire de Gériatrie de Montréal* (CRIUGM). All participants were cognitively unimpaired and overall healthy. Exclusion criteria included cognitive complaint/worry, stroke, head trauma, psychiatric disorders, and magnetic resonance imaging (MRI)-related contraindications, such as metal in the body (e.g., pacemaker), claustrophobia, and pregnancy. Ethical approval was obtained from the aging-neuroimaging research ethics committee of the CIUSSS du *Centre-Sud-de-île-de-Montréal* (CIUSSS-CSMTL), and from the McGill University Institutional Review Board, Faculty of Medicine and Health Sciences. Written informed consent was obtained from all participants.

### 2.2 Functional Testing

Three measures sensitive to processing speed, manual dexterity, and gait/mobility were acquired, namely: (a) Processing Speed Test (PST) [26], based on the Symbol-Digit Modalities Test (SDMT), measuring processing speed, (b) the 9-hole Peg Test (9HPT), assessing manual dexterity, and (c) the Timed 25-foot Walk Test (T25FW) for gait and mobility. A description of each test has been provided in Supplementary Table S1.

### 2.3 MR Acquisition

*In vivo* brain data were acquired on a 7T Siemens Terra MR scanner at the McConnell Brain Imaging Centre of the Montreal Neurological Institute-Hospital (MNI; McGill University), using an 8-channel transmit and 32-channel receive head coil. *T_1_*-weighted Magnetization-Prepared 2 Rapid Acquisition Gradient-Echo (MP2RAGE) images [27] were obtained for localization (TR/TE = 5000/2.33 ms; field of view (FOV): 240 mm; slice thickness = 0.70 mm; 1 average; 240 slices; acceleration factor = 3; acquisition time: 8 min 55 s). Using these anatomical images as reference, two MRS single-voxels were positioned (Fig. 1), one over the PCC/precuneus, a metabolically active hub region [11] (predominantly grey matter, GM), and the other over the CSWM, a region prone to age-related white matter damage [28] (predominantly white matter, WM). The PCC voxel (30 x 20 x 20 mm³) was positioned at midline aligned posterior to the corpus callosum, and the CSWM voxel (30 x 15 x 15 mm³) was placed in the left supraventricular WM. MRS spectra were acquired using the STimulated Echo Acquisition Mode (STEAM) sequence with the following parameters: TR/TE = 5000/8 ms; mixing time (TM) = 40 ms; flip angle = 90°; 64 to 96 averages in PCC, and 96 to 128 averages in CSWM; 4096 points; 5000 Hz spectral bandwidth; and acquisition time: 5 to 8 min in PCC, and 8 to 10 min in CSWM. Shimming was performed automatically using the Siemens “brain” B0 shim mode [29], and VAriable Power and Optimized Relaxations delays (VAPOR) method was used for water suppression [30]. Non-suppressed water spectra were acquired for eddy current correction and water reference quantification (TR/TE = 5000/8.00 ms; TM = 40 ms; 8 averages). Each scan lasted about 1 hour 15 minutes.

**Figure 1.**
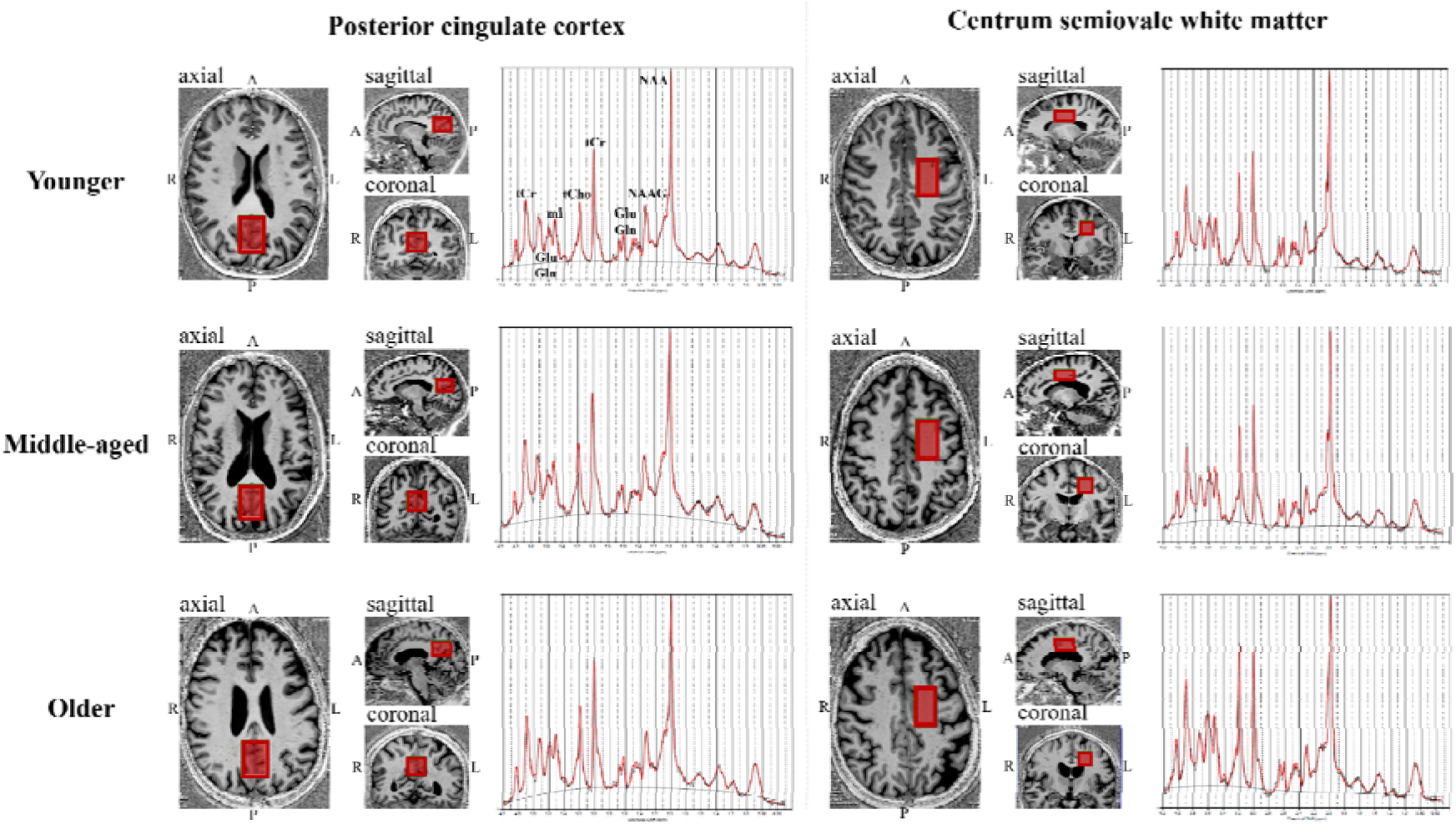
Single-voxel (red boxes) placement per age group (i.e., Younger, Middle-aged, Older) in posterior cingulate cortex/precuneus; and left centrum semiovale white matter. *Abbreviations:* Gln, glutamine; Glu, glutamate; mI, *myo*-inositol; NAA, *N*-acetylaspartate; NAAG, *N*-acetylaspartyl-glutamate; PCr, phosphocreatine; RL x AP x FH, right-left x anterior-posterior x foot-head; tCho, total choline; tCr, total creatine. Glutathione (GSH) was not labeled on the example spectrum due to its complex structure with multiple overlapping resonances.

### 2.4 Image Processing and Analysis

*T_1_*-weighted images were segmented into GM, WM, and cerebrospinal fluid (CSF) using the locally-developed Brain Tissue Composition-Net (BTCNet) pipeline, based on a published framework [31]. Each MRS voxel was converted into a binary mask image in the *T_1_*-weighted image space using another locally-developed software, built using the MINC Toolkit (https://bic-mni.github.io), and the fractions of the MRS voxel composed of GM, WM, and CSF were calculated.

### 2.5 Spectral Processing and Quantification

#### 2.5.1 MRS Preprocessing and Processing

Raw MRS data was preprocessed using the FID Appliance (FID-A) [32] in Matlab (v2022b; MathWorks Inc. Natick, USA), before spectral fitting. Eddy current correction and spectral analysis were performed in LCModel (v6.3; Provencher Inc., Canada) [33], using a basis set consisting of 21 metabolite basis spectra and 9 macromolecule basis spectra. Specifically, the following metabolites were included in the basis set: alanine (Ala), aspartate (Asp), phosphocholine (PCh), creatine (Cr), phosphocreatine (PCr), gamma-aminobutyric acid (GABA), glutamine (Gln), glutamate (Glu), glutathione (GSH), glycine (Gly), *myo*-inositol (Ins), lactate (Lac), *N*-acetylaspartate (NAA), scyllo-inositol (Scyllo), taurine (Tau), glucose (Glc), *N*-acetylaspartyl-glutamate (NAAG), glycerophosphocholine (GPC), phosphatidylethanolamine (PE), serine (Ser), ascorbate (Asc), and nine macromolecule resonances (MM1-MM9). Macromolecular frequencies were: 0.89, 1.2, 1.39, 1.63, 1.98, 2.28, 2.98, 3.19, and 3.75 ppm.

#### 2.5.2 Absolute Quantification

Following expert consensus recommendations [34], quantification of metabolite signal intensity to absolute concentrations was performed using the equation (Eq. 1; Supplementary Material S1) provided by Dhamala et al. [35]. *T_1_* and *T_2_* relaxation time constants of water at 7T were obtained from the literature [36,37]: *T_1_* (WM) = 1120 ms; *T_2_* (WM) = 55 ms; *T_1_* (GM) = 2130 ms; *T_2_* (GM) = 50 ms; *T_1_* (CSF) = 4425 ms; *T_2_* (CSF) = 141 ms. Assumed visible water concentration was 36100 mmol/L in WM, 43300 mmol/L in GM, and 53800 mmol/L in CSF [35]. Voxel-specific tissue water concentrations were calculated per participant using Eq. 2 (Supplementary Material S1). Given that our TR >> TE, the *T_2_* relaxation times of metabolites, but not *T_1_*, were included from the literature [38] (see Supplementary Table S2). Finally, GM, WM, and CSF volume fractions within the two MRS voxels were determined from structural data, and ranges are provided in Supplementary Table S3.

#### 2.5.3 Quality Control

Data quality control was performed via visual inspection of the spectrum, fit, baseline, and residual, as per recommendation [39]. Signal-to-Noise Ratio (SNR) and Full-Width at Half Maximum (FWHM) were also inspected for each spectrum. Metabolites where the Cramér-Rao Lower Bound (CRLB) estimates (as provided by LCModel), averaged across all participants, exceeded 20% were considered unreliable and systematically excluded, following a commonly used threshold [6]. The resulting mean CRLBs for included metabolites per region of interest are shown in Supplementary Table S4.

### 2.6 Statistical Analysis

All statistical analyses were conducted in R (v4.4.1; R Core Team) within RStudio (v2024.09.0; RStudio, PBC, USA). General linear models (GLMs) were used to assess the relationship between metabolite levels and (a) age and (b) age groups, controlling for sex. To assess the effect of sex on age-related metabolite changes, participants were divided into two groups (i.e., men and women), and the associations with age and age group were performed independently within each sex. To investigate whether menopausal status impacts age-related metabolite level variations, women were categorized based on self-reported pre- and post-menopausal status, with an age cut-off of 50 years of age (i.e., women < 50 years of age: pre-menopausal; women > 50 years of age: post-menopausal). Furthermore, to control for the peri-menopausal transition phase and self-reported variability, our final menopausal status groups were based on ± 5 years of the age cut-off, resulting in pre-menopausal women younger than 45 years of age, and post-menopausal women aged 55+ years of age. These ages were consistent with previously reported menopausal age ranges [40,41]. Additionally, to control for the effect of age in menopausal associations, the same analysis was performed in age-matched men (< 45 vs. > 55 years of age). Finally, GLMs, controlling for age, were performed to assess the effect of metabolite levels on functional test scores, in all participants, and in men and women separately. All statistical analyses were corrected for multiple comparisons using the false discovery rate (FDR) method at 5%. Statistical significance threshold was set at *p* < 0.05. Metabolite concentrations are reported in institutional units.

## 3. RESULTS

Participant demographics are provided in Table 1. Age groups were divided as follows: young adults: 20-39 years of age; middle-aged adults: 40-59 years of age; and old adults: 60-79 years of age. After performing quality control, five participants were excluded for analyses for PCC (four men and one woman in the older group) and one for CSWM (man in the younger group), resulting in 90 participants for PCC and 94 for CSWM. For all participants, there was 100% concordance between sex (biological) and gender (social), therefore the term “sex” is being used to describe both biological and social variables. All results are summarized in Table 2.

**Table 1.**
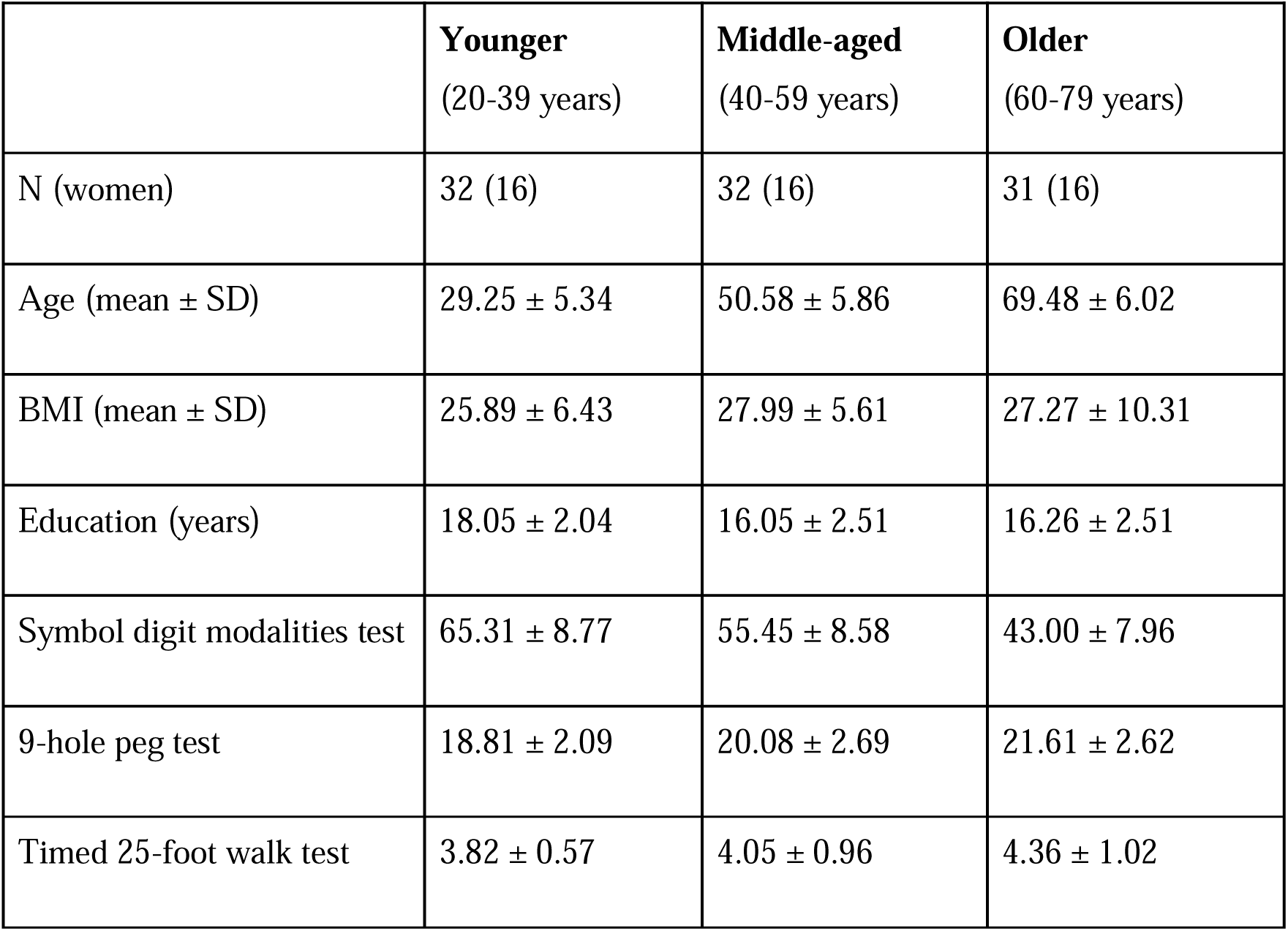
Participant demographics. *Abbreviations:* BMI, body-mass index = weight (kg) / [height (m)] ²; SD, standard deviation.

|  | **Younger**<br>(20-39 years) | **Middle-aged**<br>(40-59 years) | **Older**<br>(60-79 years) |
| --- | --- | --- | --- |
| N (women) | 32 (16) | 32 (16) | 31 (16) |
| Age (mean ± SD) | 29.25 ± 5.34 | 50.58 ± 5.86 | 69.48 ± 6.02 |
| BMI (mean ± SD) | 25.89 ± 6.43 | 27.99 ± 5.61 | 27.27 ± 10.31 |
| Education (years) | 18.05 ± 2.04 | 16.05 ± 2.51 | 16.26 ± 2.51 |
| Symbol digit modalities test | 65.31 ± 8.77 | 55.45 ± 8.58 | 43.00 ± 7.96 |
| 9-hole peg test | 18.81 ± 2.09 | 20.08 ± 2.69 | 21.61 ± 2.62 |
| Timed 25-foot walk test | 3.82 ± 0.57 | 4.05 ± 0.96 | 4.36 ± 1.02 |

**Table 2.**
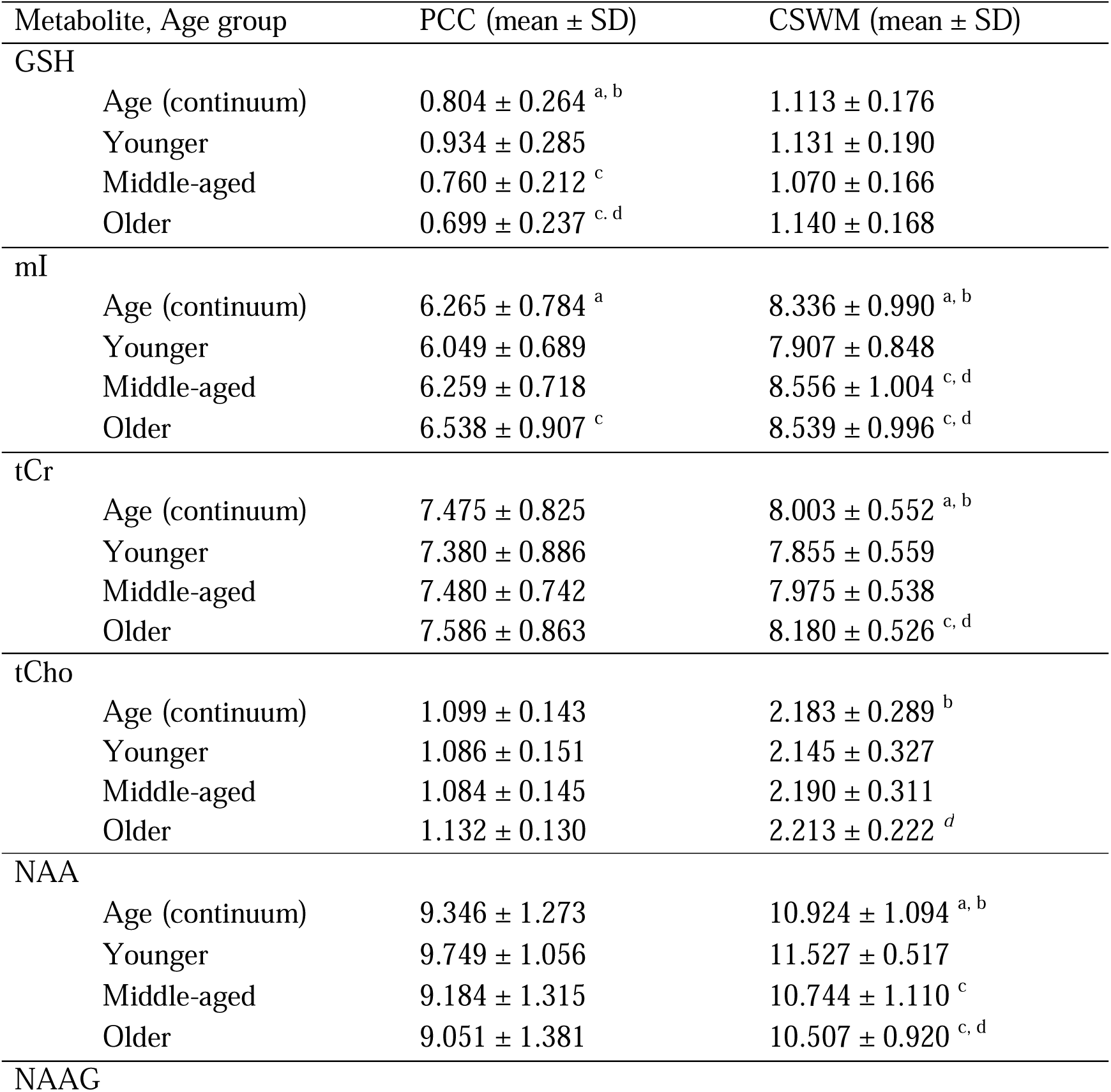
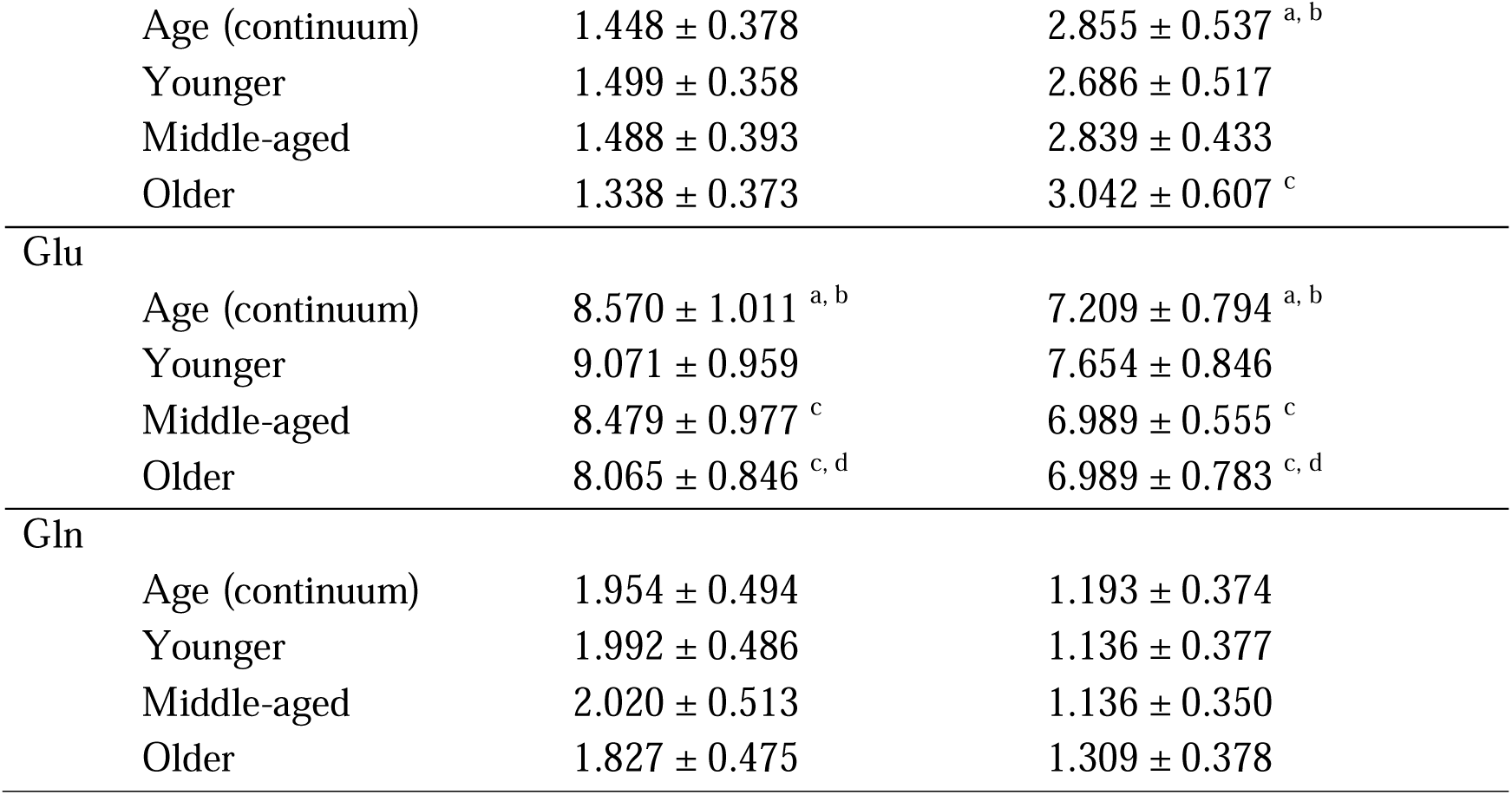
Metabolite levels per region of interest, and statistical significance with age (continuum and groups) and sex. *Abbreviations:* ^a^, significant association with age as a continuum; ^b^, significant association with age as a continuum in women only; ^c^, significant group differences relative to younger adults; CSWM, centrum semiovale white matter; ^d^, significant group differences relative to younger adults in women only; Gln, glutamine; Glu, glutamate; GSH, glutathione; *italics,* borderline significance; mI, *myo*-inositol; NAA, *N*-acetylaspartate; NAAG, *N*-acetylaspartyl-glutamate; PCC, posterior cingulate cortex; tCho, total choline; tCr, total creatine.

| Metabolite, Age group | PCC (mean ± SD) | CSWM (mean ± SD) |
| --- | --- | --- |
| **GSH** |  |  |
| Age (continuum) | 0.804 ± 0.264 <sup>a, b</sup> | 1.113 ± 0.176 |
| Younger | 0.934 ± 0.285 | 1.131 ± 0.190 |
| Middle-aged | 0.760 ± 0.212 <sup>c</sup> | 1.070 ± 0.166 |
| Older | 0.699 ± 0.237 <sup>c, d</sup> | 1.140 ± 0.168 |
| **mI** |  |  |
| Age (continuum) | 6.265 ± 0.784 <sup>a</sup> | 8.336 ± 0.990 <sup>a, b</sup> |
| Younger | 6.049 ± 0.689 | 7.907 ± 0.848 |
| Middle-aged | 6.259 ± 0.718 | 8.556 ± 1.004 <sup>c, d</sup> |
| Older | 6.538 ± 0.907 <sup>c</sup> | 8.539 ± 0.996 <sup>c, d</sup> |
| **tCr** |  |  |
| Age (continuum) | 7.475 ± 0.825 | 8.003 ± 0.552 <sup>a, b</sup> |
| Younger | 7.380 ± 0.886 | 7.855 ± 0.559 |
| Middle-aged | 7.480 ± 0.742 | 7.975 ± 0.538 |
| Older | 7.586 ± 0.863 | 8.180 ± 0.526 <sup>c, d</sup> |
| **tCho** |  |  |
| Age (continuum) | 1.099 ± 0.143 | 2.183 ± 0.289 <sup>b</sup> |
| Younger | 1.086 ± 0.151 | 2.145 ± 0.327 |
| Middle-aged | 1.084 ± 0.145 | 2.190 ± 0.311 |
| Older | 1.132 ± 0.130 | 2.213 ± 0.222 <sup>d</sup> |
| **NAA** |  |  |
| Age (continuum) | 9.346 ± 1.273 | 10.924 ± 1.094 <sup>a, b</sup> |
| Younger | 9.749 ± 1.056 | 11.527 ± 0.517 |
| Middle-aged | 9.184 ± 1.315 | 10.744 ± 1.110 <sup>c</sup> |
| Older | 9.051 ± 1.381 | 10.507 ± 0.920 <sup>c, d</sup> |
| **NAAG** |  |  |

|  |  |  |  |
| --- | --- | --- | --- |
| | Age (continuum) | $1.448 \pm 0.378$ | $2.855 \pm 0.537^{a, b}$ |
| | Younger | $1.499 \pm 0.358$ | $2.686 \pm 0.517$ |
| | Middle-aged | $1.488 \pm 0.393$ | $2.839 \pm 0.433$ |
| | Older | $1.338 \pm 0.373$ | $3.042 \pm 0.607^c$ |
| Glu | Age (continuum) | $8.570 \pm 1.011^{a, b}$ | $7.209 \pm 0.794^{a, b}$ |
| | Younger | $9.071 \pm 0.959$ | $7.654 \pm 0.846$ |
| | Middle-aged | $8.479 \pm 0.977^c$ | $6.989 \pm 0.555^c$ |
| | Older | $8.065 \pm 0.846^{c, d}$ | $6.989 \pm 0.783^{c, d}$ |
| Gln | Age (continuum) | $1.954 \pm 0.494$ | $1.193 \pm 0.374$ |
| | Younger | $1.992 \pm 0.486$ | $1.136 \pm 0.377$ |
| | Middle-aged | $2.020 \pm 0.513$ | $1.136 \pm 0.350$ |
| | Older | $1.827 \pm 0.475$ | $1.309 \pm 0.378$ |

### 3.1 Age-Specific Metabolite Level Changes

#### 3.1.1 Age as a Continuum

After controlling for sex, increasing age (20-79 years; Fig. 2) was associated with higher mI levels and lower Glu levels and in both PCC (mI: *p* = 0.005; Glu: *p* = 0.003) and CSWM (mI: *p* = 0.005; Glu: *p* = 0.003). In PCC, we found lower GSH levels (*p* = 0.003) with increasing age. Furthermore, in CSWM, increasing age was associated with (b) higher tCr (*p* = 0.009), (b) lower NAA (*p* = 0.003), and (c) higher NAAG (*p* = 0.008) levels.

**Figure 2.**
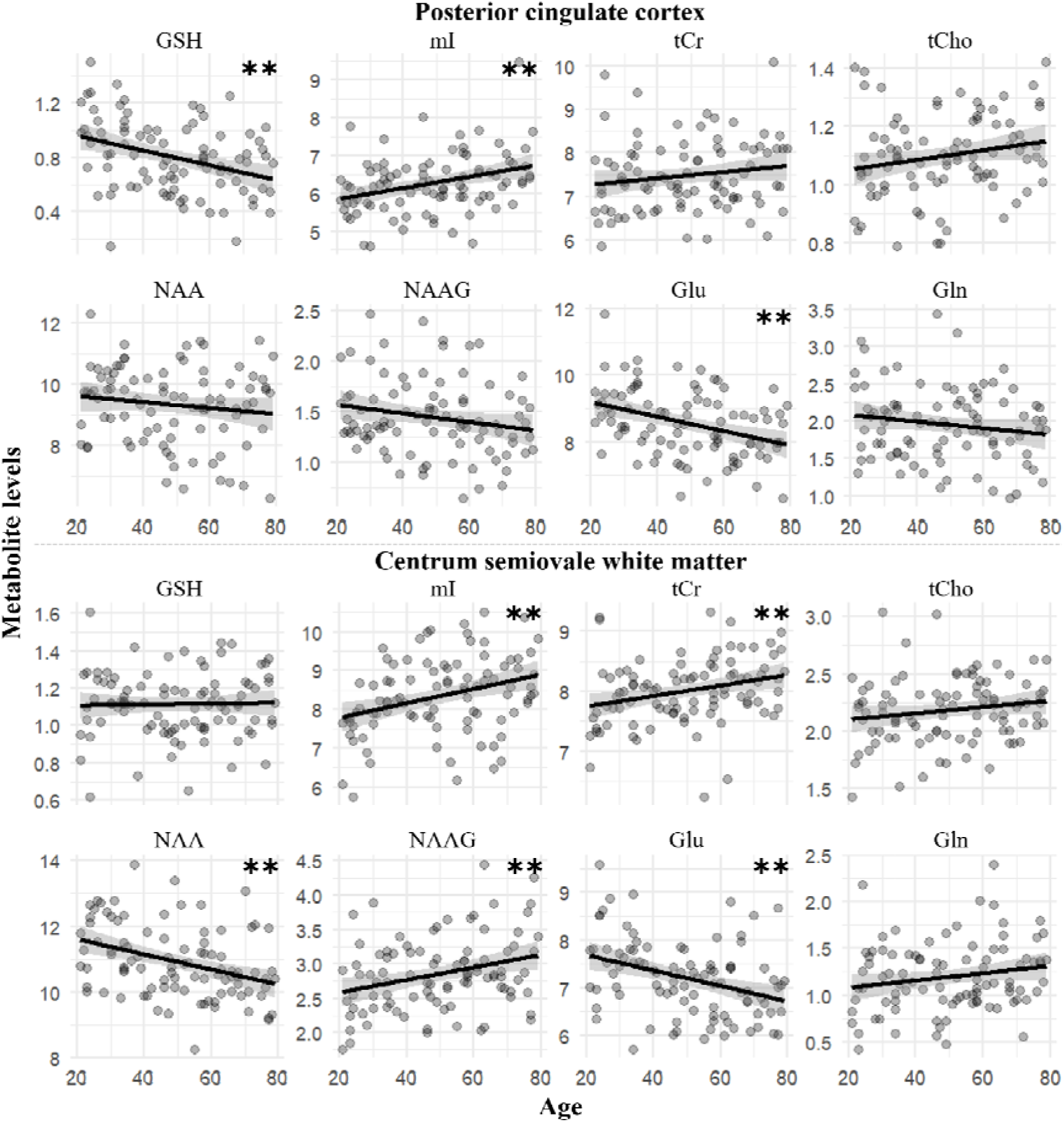
Age-specific metabolite level changes with age as a continuum. *Abbreviations:* **, *p* < 0.01; Gln, glutamine; Glu, glutamate; GSH, glutathione; mI, *myo*-inositol; NAA, *N*-acetylaspartate; NAAG, *N*-acetylaspartyl-glutamate; tCho, total choline; tCr, total creatine.

#### 3.1.2 Age Groups

When investigating metabolite level changes by age group (i.e., Middle-aged vs. Younger, Older vs. Younger, and Older vs. Middle-aged), we found similar results (Fig. 3). Specifically, in both PCC and CSWM, we found (a) higher mI levels (PCC: *p* = 0.044; CSWM: *p* = 0.032) in older adults relative to younger adults, and (b) lower Glu levels in middle-aged (PCC: *p* = 0.049; CSWM: *p* = 0.010) and older (PCC: *p* = 0.001; CSWM: *p* = 0.003) adults relative to younger adults. In addition, in PCC, we found lower GSH levels in middle-aged (*p* = 0.036) and older (*p* = 0.003) adults compared to younger adults. In CSWM, relative younger adults, (a) middle-aged adults had higher levels of mI (*p* = 0.036) and lower levels of NAA (*p* = 0.026), and (b) older adults had higher tCr (*p* = 0.042), lower NAA (*p* = 0.001), and higher NAAG (*p* = 0.029) levels.

**Figure 3.**
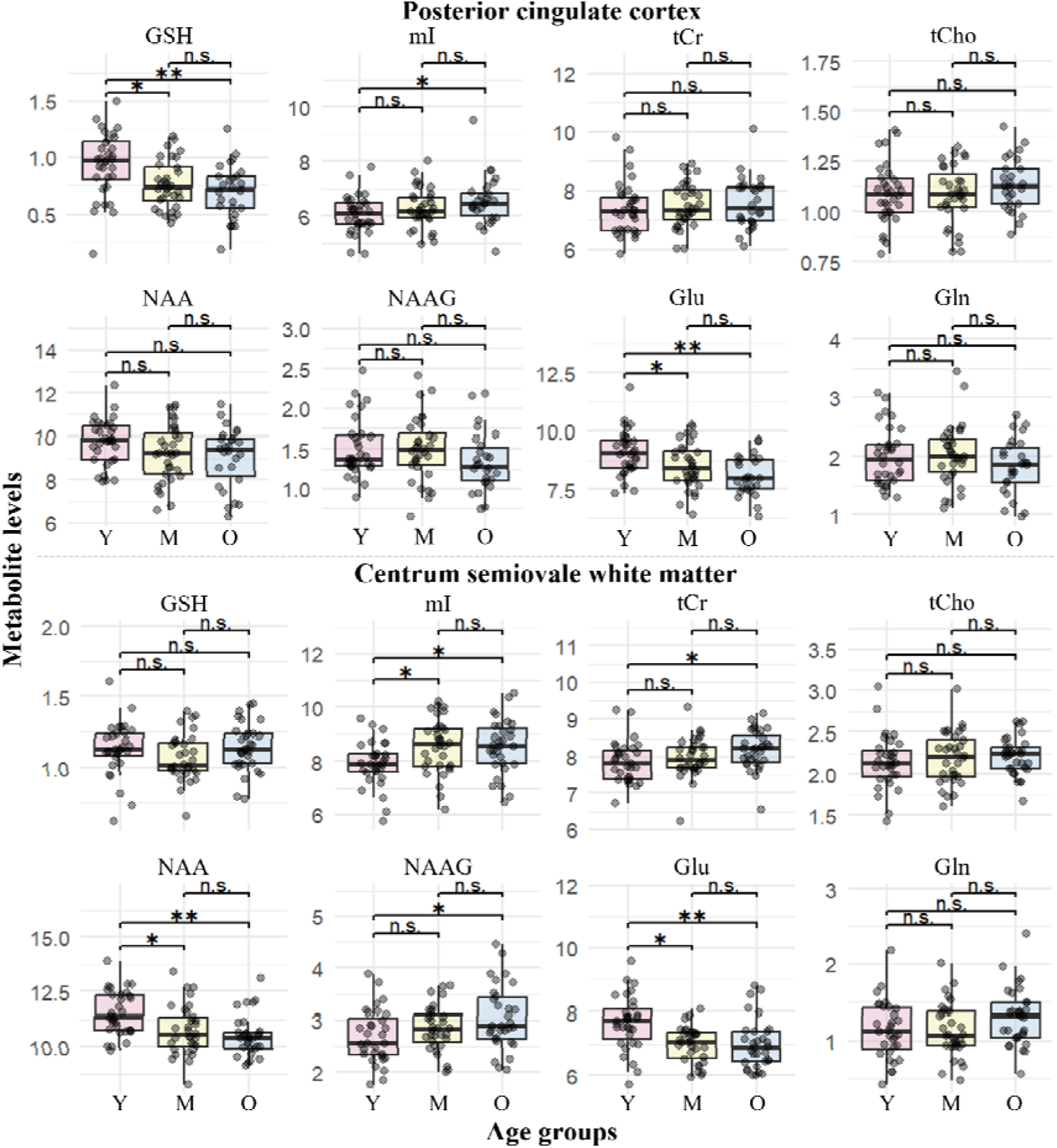
Age-specific metabolite level changes between age groups. *Abbreviations:* *, *p* < 0.05; **, *p* < 0.01; Gln, glutamine; Glu, glutamate; GSH, glutathione; M, middle-aged; mI, *myo*-inositol; NAA, *N*-acetylaspartate; NAAG, *N*-acetylaspartyl-glutamate; n.s., not significant; O, older; tCho, total choline; tCr, total creatine; Y, younger.

### 3.2 Sex-Specific Metabolite Level Changes

When investigating men and women as two independent groups (Fig. 4), we found that findings were driven by age-related changes in women.

**Figure 4.**
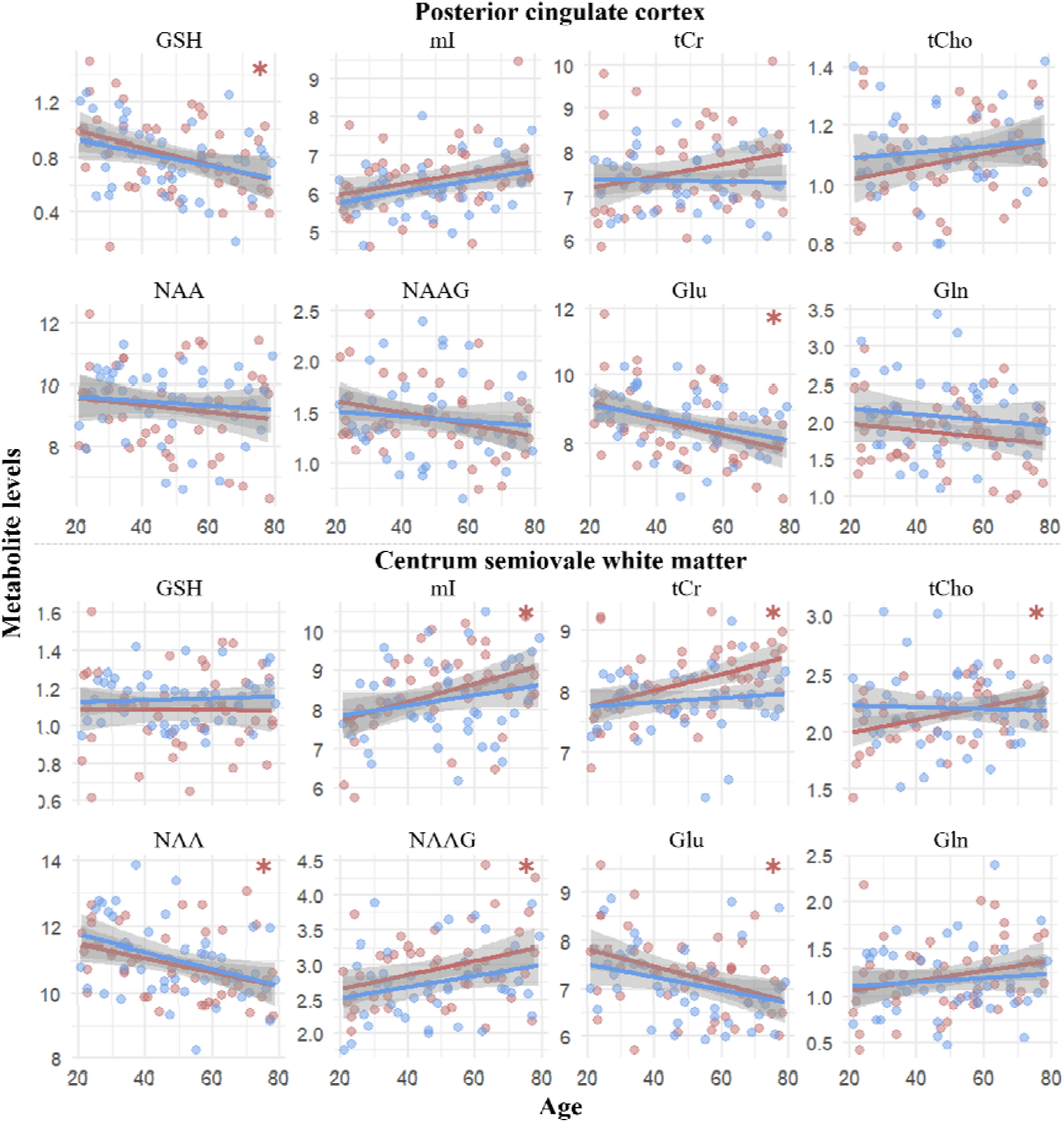
Sex-specific metabolite level changes with increasing age. Brown represents women and blue represents men. *Abbreviations:* *, *p* < 0.05; Gln, glutamine; Glu, glutamate; GSH, glutathione; mI, *myo*-inositol; NAA, *N*-acetylaspartate; NAAG, *N*-acetylaspartyl-glutamate; tCho, total choline; tCr, total creatine.

#### 3.2.1 Age as a Continuum

With increasing age, men did not demonstrate any significant metabolite level changes. In contrast, in women, increasing age was associated with lower Glu levels in both PCC (*p* = 0.022) and CSWM (*p* = 0.020). In PCC, women demonstrated lower GSH levels (*p* = 0.022) with increasing age. Finally, in CSWM, increasing age in women was associated with (a) higher levels of mI (*p* = 0.018), tCr (*p* = 0.018), tCho (*p* = 0.018), and NAAG (*p* = 0.040), and (b) lower NAA levels (*p* = 0.024).

#### 3.2.2 Age Groups

Within-sex analyses showed no significant differences in metabolite levels across age groups in men. In women, age group effects were pronounced. Older women had significantly lower Glu levels in PCC (*p* = 0.019) and CSWM (*p* = 0.024) than younger women. In addition, in PCC, GSH levels were lower in older relative to younger women (*p* = 0.024). In CSWM, (a) middle-aged women had higher mI (*p* = 0.011) levels relative to younger women, and (b) older women had higher levels of mI (*p* = 0.024) and tCr (*p* = 0.024), and lower NAA levels (*p* = 0.024) compared to younger women. Finally, higher tCho levels in older women compared to younger women were borderline significant in CSWM (*p* = 0.054).

#### 3.2.3 Menopausal Status

To investigate whether metabolite level changes differed by menopausal status, women were stratified based on their self-reported status (± 5 years to control for those undergoing a peri-menopausal transition phase). In particular, 19 women were pre-menopaused (30.53 ± 7.19 years of age) and 21 were post-menopaused (66.71 ± 7.31 years of age). In particular, we found that, relative to pre-menopausal women, post-menopausal women had (a) lower Glu levels in PCC (*p_unadjusted_* = 0.022) and CSWM (*p_unadjusted_* = 0.015), (b) higher tCho levels in PCC (*p_unadjusted_* = 0.044) and CSWM (*p_unadjusted_* = 0.006), (c) lower GSH levels (*p_unadjusted_* = 0.024) in PCC, and (c) higher levels of mI (*p_unadjusted_* = 0.014) and tCr (*p_unadjusted_*= 0.004) in CSWM, but none of these results survived multiple comparisons correction. Notably, with multiple comparisons correction, differences in tCho and tCr levels in CSWM between pre-and post-menopausal women were borderline significant (*p* = 0.058).

When comparing men older than 55 years (N = 19; 67.00 ± 7.37 years of age) relative to those younger than 45 years (N = 18; 31.33 ± 6.02 years of age), we found that older men had (a) lower Glu levels in PCC (*p_unadjusted_* = 0.013) and CSWM (*p_unadjusted_* = 0.045), (b) lower GSH levels (*p_unadjusted_*= 0.043) and higher mI levels (*p_unadjusted_* = 0.011) levels in PCC, and (c) lower NAA levels (*p_unadjusted_* = 0.010) in CSWM, but none of these results survived multiple comparisons correction.

### 3.3 Metabolite Levels and Functional Test Scores

No association between metabolite levels and functional test scores remained significant after correcting for multiple comparisons correction. Without correction, we found a positive association between 9HPT score and tCho levels in PCC (*p_unadjusted_*= 0.033). In women, 9HPT score was negatively associated with mI (*p_unadjusted_*= 0.048) and NAAG (*p_unadjusted_* = 0.006) levels in CSWM. Finally, in men, we found positive associations between (a) SDMT score and NAA levels in CSWM (*p_unadjusted_* = 0.047), and (b) 9HPT score and Gln levels in CSWM (*p_unadjusted_* = 0.037).

With increasing age, there was a negative association with the SDMT score (*p* < 0.001), and positive associations with the T25FW (*p* = 0.029) and 9HPT (*p* < 0.001) scores. Specifically, middle-aged (*p* < 0.001) and older adults (*p* < 0.001) had lower SDMT scores (i.e., worse processing speed) than younger adults, and older adults (*p* < 0.001) also had lower scores compared to middle-aged adults. In addition, (a) older adults had a higher T25FW score (i.e., longer time to walk 25 feet) than younger adults (*p* = 0.015), (b) older adults had a higher 9HPT score (i.e., worse manual dexterity) relative to both middle-aged (*p* = 0.017) and younger (*p* < 0.001) adults, and (c) middle-aged adults had a higher 9HPT score compared to younger adults (*p* = 0.044). Sex-specific analyses revealed that age was (a) negatively associated with the SMDT score (women: *p* < 0.001; men: *p* < 0.001) and (b) positively associated with the 9HPT score (women: *p* = 0.002; men: *p* = 0.001).

## 4. DISCUSSION

Our study examines the healthy adult lifespan, from younger to older age, with particular focus on midlife, a period often underrepresented in research but critical for future health [2,3]. Using state-of-the-art UHF MRS with expert consensus methods [34,39], we precisely quantified eight metabolites, including three low-concentration metabolites, revealing age-related metabolic changes in the brain. These metabolic pathways in different brain cell types have been summarized in Figure 5. Our results indicate that early signatures of OS, neuroinflammation, and neuronal vulnerability emerge in midlife, reflected by changes in (a) GSH, (b) mI, tCr, and tCho, and (c) NAA, NAAG, and Glu levels, respectively. These interconnected processes appear fundamental to brain aging, and may contribute to onset and progression of NDDs such as AD [7,42,43]. Importantly, they align with current therapeutic priorities, as evidenced by the 2025 AD drug development pipeline, which includes 31% of agents targeting metabolism, OS, neuroinflammation, and neuronal processes [44].

**Figure 5.**
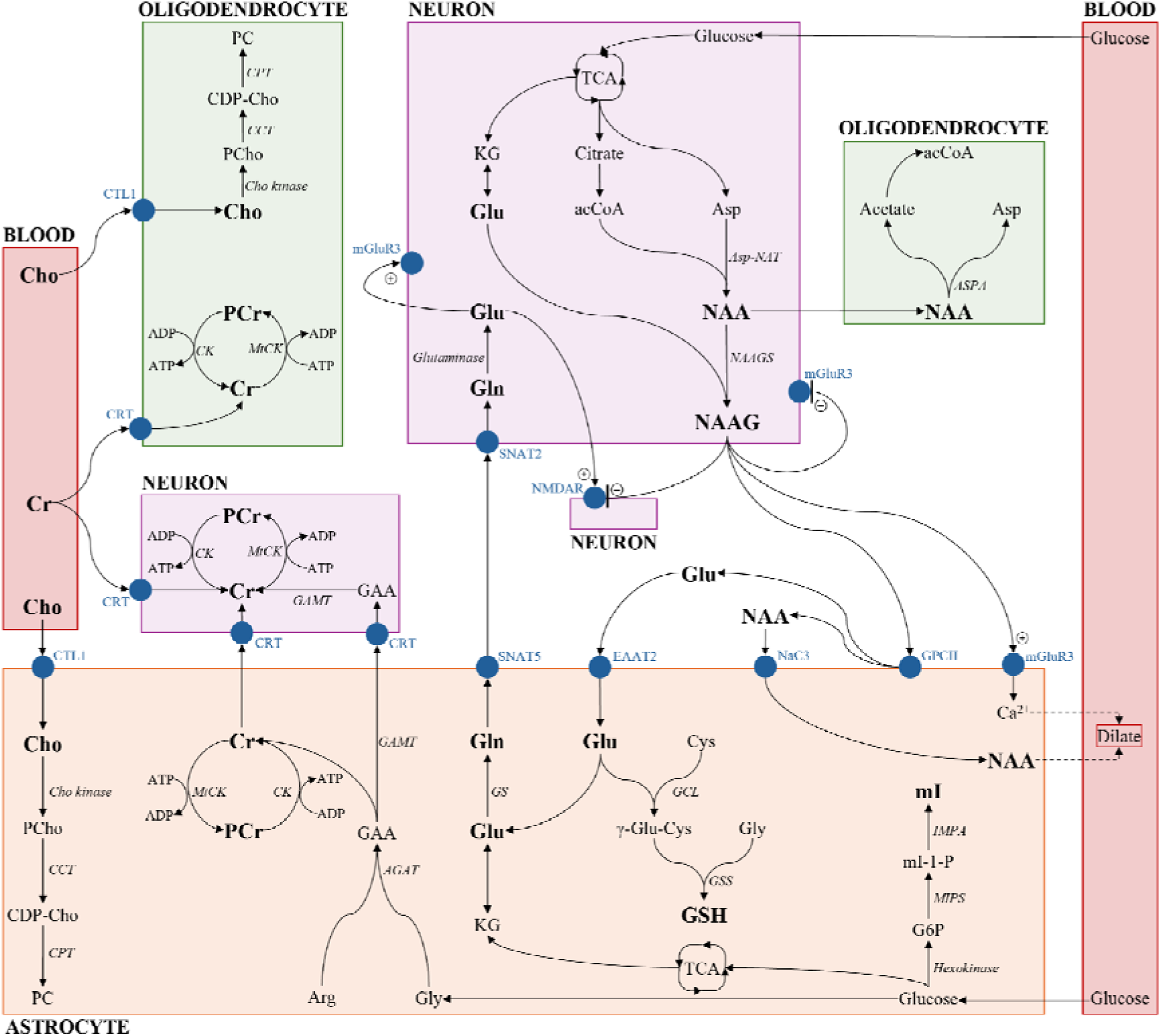
Depiction of metabolic pathways in different brain cell types. Our MRS-detected metabolites are in bold. Enzymes are in italics. Transporters and receptors are in blue. *Abbreviations: +*, stimulation; -, inhibition; acCoA, acetyl coA; AGAT, arginine-glycine amidinotransferase; Arg, arginine; Asp, aspartate; Asp-NAT, aspartate N-acetyltransferase; ASPA, aspartoacylase; ADP, adenosine diphosphate; ATP, adenosine triphosphate; Ca^2+^, calcium; CCT, phosphocholine cytidylyltransferase; CDP-Cho, cytidine diphosphate-choline; Cho, choline; CK, creatine kinase; CPT, cholinephosphotransferase; Cr, creatine; CRT, creatine transporter; CTL1, choline transporter-like 1 protein; Cys, cysteine; EAAT2, excitatory amino acid transporter 2; G6P, glucose-6-phosphate; GAA, guanidinoacetate; GAMT, guanidinoacetate methyltransferase; GCL, glutamate-cysteine ligase; Gln, glutamine; Glu, glutamate; GPCII, glutamate carboxypeptidate II receptor; GSH, glutathione; GS, glutamine synthetase; GSS, glutathione synthetase; Gly, glycine; γ-Glu-Cys, gamma-glutamate-cysteine; IMPA, inositol monophosphate; KG, α-ketoglutarate; mGluR3, metabotropic Glu receptor 3; mI, *myo*-inositol; mI-1-P, myo-inositol-1-phosphate; MIPS, mI phosphate synthase; mtCK, mitochondrial CK; NAA, *N*-acetylaspartate; NAAG, *N*-acetylaspartyl-glutamate; NAAGS, NAAG synthase; NMDAR, *N*-methyl-D-aspartate receptor; PC, phosphatidylcholine; PCr, phosphocreatine; PCho, phosphocholine; SNAT2, amino acid transporter 2; SNAT5, amino acid transporter 5; TCA, tricarboxylic acid cycle.

### 4.1 Age-Related Metabolite Changes in Healthy Aging

#### 4.1.1 Midlife Antioxidant Decline in Healthy Aging

In our cross-sectional UHF MRS study, levels of the antioxidant GSH in the PCC showed a negative association with age as a continuous measure, and age-group comparisons demonstrated reduced concentrations by midlife, highlighting early vulnerability to OS. Parallel to our findings, a prior 7T MRS study reported a negative association between GSH levels and age in the anterior cingulate cortex in healthy adults (20-54 years), although midlife-specific changes were not assessed [45]. Although MRS studies measuring GSH remain limited due to its complex structure complicating quantification at lower field strengths [7], these observations suggest that OS in GM may increase earlier in adulthood than previously recognized.

This vulnerability is reinforced by the high metabolic demand of GM, which consumes over twice the oxygen of WM and contains more mitochondria to support adenosine triphosphate (ATP) production via oxidative phosphorylation – the predominant ROS source [5,46]. Elevated energetic burden renders GM, and particularly highly-connected hub regions like the PCC [11,12,23], prone to OS, which is further amplified by the midlife GSH decline observed in our study. While modest OS is typical with aging, excessive elevations may signal early neuropathological risk [5], consistent with evidence of reduced antioxidant capacity in PCC in AD [47]. Measuring GSH in PCC using UHF MRS could therefore provide an early marker of abnormal metabolic shifts and a targetable window for intervention [7,48].

#### 4.1.2 Midlife Neuroinflammation in Healthy Aging

Importantly, OS in midlife may act in concert with neuroinflammatory processes, further contributing to early brain vulnerability. In our study, age-group comparisons revealed elevated mI levels in midlife in CSWM, while continuous age analyses showed generally higher mI with increasing age in both PCC and CSWM. Only one prior 7T MRS study included middle-aged adults and reported higher mI levels without directly assessing WM [49], highlighting the novelty of our reported higher WM mI in midlife. Our findings likely reflect early WM neuroinflammation, as mI is predominantly localized in glial cells and its elevation has been linked to astrogliosis [13] and systemic inflammation [49,50], supporting its interpretation as an *in vivo* marker of neuroimmune activity. This interpretation is further reinforced by complementary evidence from postmortem immunohistochemistry of brain tissue and *in vivo* PET imaging using [¹¹C]-(R)-PK11195 – a ligand targeting activated microglia – both demonstrating increased WM neuroimmune activation beginning in midlife [51]. In NDDs, including AD, WM neuroinflammation has been reported to precede symptom onset [52], and increased MRS-detected mI levels have been found early in the AD continuum [53], highlighting its potential role as a biomarker of neurodegenerative processes.

Similar to mI, tCr is higher in glial cells than neurons, and has been associated with neuroinflammation [49,54]. We observed age-related increases in tCr in CSWM, with significant differences between older and younger adults. This pattern aligns with most MRS studies reporting tCr elevations with age in both PCC and CSWM [15,16,55,56]. Since creatine exhibits antioxidant and anti-inflammatory properties [57], these increases in healthy aging may reflect a compensatory mechanism to altered energy demands [56]. By contrast, reduced tCr levels in AD [58] could indicate a breakdown of this compensatory mechanism, along with overall reduced metabolic activity due to neurodegeneration.

Together, these findings suggest that both mI and tCr serve as sensitive biomarkers of early neuroinflammatory changes, potentially heralding vulnerability to neurodegeneration in AD.

#### 4.1.3 Midlife Neuronal Vulnerability in Healthy Aging

OS and neuroinflammation can both contribute to neurodegeneration and NDDs [42]. In CSWM, we observed lower NAA levels with increasing age, with age-group comparisons showing decreases starting in middle-aged adults. Given that NAA is an established neuroaxonal integrity marker [13], our results point to emerging neuroaxonal impairment beginning in midlife. This interpretation is supported by a study combining MRS and diffusion tensor imaging in middle-aged-to-older adults demonstrating WM ultrastructure alterations [59], although younger adults and age-group differences were not examined. In most NDDs, including AD, decreased NAA levels have been reported, reflective of neuronal dysfunction and/or loss and mitochondrial dysfunction [58,60].

In contrast to NAA, literature on the neuromodulator NAAG is sparse, given its much lower concentration and quantification complexity (especially at lower field strengths), which often leads to its combination with NAA as total NAA [15,16]. We found higher levels of NAAG in CSWM with increasing age. Only one 7T study assessed NAAG levels and found decreases in older relative to younger adults [22], without investigating WM or middle-aged adults, highlighting the novelty of our finding. High NAAG levels are considered neuroprotective and have been found to have a positive effect on cognition [61]. It also modulates Glu release, therefore preventing Glu-induced excitotoxicity [61]. Given its action on Glu receptors and decreased levels reported in several NDDs, including AD, NAAG has become a focus of clinical trials [61,62].

In fact, our study provides novel evidence that Glu levels decrease in the PCC and CSWM starting in midlife. Prior 7T MRS studies observed lower Glu only in GM and at older ages, leaving midlife unexplored [22,49]. In PCC, lower midlife Glu levels likely reflect disruptions in the neuronal-astrocytic Glu-Gln cycle, involving several proteins, transporters, and receptors, leading to impaired Glu homeostasis and vulnerability to neurotoxicity [63,64]. In contrast, in CSWM, lower Glu levels may reflect compromised neuroaxonal integrity and neuroinflammation, which is supported by our findings of lower NAA levels and higher mI and tCr levels, respectively. While these changes reflect normal aging, Glu levels decline further in AD, indicating progressive metabolic and neuronal compromise linked to memory impairment [65].

Overall, these findings suggest that NAA and Glu may serve as biomarkers of increased neuronal vulnerability, while NAAG may be linked to neuroprotection in healthy aging and possibly provide a therapeutic target in AD.

### 4.2 Sex-Related Metabolite Changes in Healthy Aging

Our sex-specific analyses revealed that the observed metabolite changes were driven primarily by women, with no significant effects in men. In women, increasing age was associated with lower GSH levels in PCC, indicating reduced antioxidant capacity, and lower Glu levels, reflecting increased neuronal vulnerability. In CSWM, increasing age corresponded to higher levels of mI, tCr, and tCho, indicative of elevated neuroinflammation, alongside lower Glu and NAA levels, suggesting greater WM compromise.

These age-related sex-specific differences in metabolite levels might be due to estrogen and other estrogenic compounds. In particular, estrogen possesses antioxidant and anti-inflammatory properties, by protecting mitochondrial function [66] and lowering pro-inflammatory markers [67], respectively. Estrogen also protects neuronal integrity by reducing Glu-induced excitotoxicity [68] and increasing NMDA availability [69]. However, this estrogen-related advantage is lost after menopause [70], typically occurring between 45-55 years of age [40,41]. Therefore, post-menopausal estrogen loss might accelerate OS, enhance neuroinflammation, and alter neuronal systems [71], overall contributing to the greater AD risk in women [72].

### 4.3 Limitations

Our study has potential limitations. First, while using state-of-the-art scanning techniques, we used two single-voxels only, due to the long MRS acquisition duration and participants’ ability/willingness to remain still in the scanner. We used a single-voxel technique rather than spectroscopic imaging in order to have sufficient spectral quality to reliably measure metabolites with low concentrations and J-coupled resonances, such as GSH. Secondly, for absolute quantification of metabolite signal intensity, similar to other studies [34,35], relaxation times were acquired from the literature rather than measured, due to scan time limitations. Moreover, while age-related decreases of *T_2_*relaxation times for NAA, tCr, and tCho have been reported [73], we used one *T_2_* value per metabolite for all participants to minimize bias, as very few studies reported *T_2_*acquired at 7T [36–38] and none covered all three age groups (especially middle-aged). Finally, we did not have biological tests available to assess hormonal levels, only self-reported menopausal status, emphasizing the need for sex-specific studies with standardized staging (e.g., Stages of Reproductive Aging Workshop guidelines [74]).

## 5. CONCLUSION

In conclusion, our cross-sectional study using state-of-the-art UHF MRS offers new insights into normative metabolite data across the healthy adult lifespan, including midlife and sex-specific patterns. In particular, we reported that OS, neuroinflammation, and neuronal vulnerability emerge as early as healthy midlife, highlighting a potentially pivotal window for brain health. In addition, our study provides evidence of sex-specific metabolic aging, with a potential sex hormone-related advantage of women (e.g., estrogen) lost after menopause, therefore contributing to greater OS, neuroinflammation, and neuronal vulnerability. Future studies should focus more on middle-aged healthy adults and the transition from pre- to post-menopause in middle-aged women, to better understand the mechanisms underlying sex-specific brain aging.

## Data Availability

All data produced in the present work are contained in the manuscript.

## ACKNOWLEDGMENTS

The authors would like to thank Ali Filali-Mouhim, PhD, from the Centre de Recherche de l’Institut Universitaire de Gériatrie de Montréal (CRIUGM) for his input in the statistical analysis, and Lianne Trigiani, PhD, for reading and providing feedback on the manuscript. The authors would also like to thank all the participants in this study for their time.

## CONSENT STATEMENT

Ethical approval was obtained from the aging-neuroimaging research ethics committee of the *CIUSSS du Centre-Sud-de-île-de-Montréal* (CIUSSS-CSMTL), and from the McGill University Institutional Review Board, Faculty of Medicine and Health Sciences. Written informed consent was obtained from all participants.

