## Supplementary Material for "Oxidative Stress, Neuroinflammation, and Neuronal Vulnerability Begin in Midlife: a 7 Tesla Magnetic Resonance Spectroscopy Healthy Adult Lifespan Study"

**Supplementary Material S1.** Equations for absolute quantification.

**Equation 1.**

$$C_{abs\_met} = \frac{LCMoutput_{met}}{conc_{H2O} \cdot ATT_{H2O}} \cdot \frac{\left(H2O_{WM} \cdot e^{-\frac{TE}{T2_{WM}}} \cdot \left(1 - e^{-\frac{TR}{T1_{WM}}}\right) \cdot f_{WM} + H2O_{GM} \cdot e^{-\frac{TE}{T2_{GM}}} \cdot \left(1 - e^{-\frac{TR}{T1_{GM}}}\right) \cdot f_{GM} + H2O_{CSF} \cdot e^{-\frac{TE}{T2_{CSF}}} \cdot \left(1 - e^{-\frac{TR}{T1_{CSF}}}\right) \cdot f_{CSF}\right)}{e^{-\frac{TE}{T2_{met}}} \cdot (f_{WM} + f_{GM})}$$

<u>With:</u>

$C_{abs\_met}$: Absolute concentration for metabolite of interest.

$LCMoutput_{met}$: LCModel output for metabolite of interest.

$ATT_{H2O} = 1$. Attenuation factor provided by LCModel (unitless).

$H2O_x$: Assumed visible water concentration for x tissue of interest (WM, GM, CSF) (mmol/L).

$T_{2\_x}$: $T_2$ relaxation time for x tissue of interest (WM, GM, CSF) (ms).

TE = 8 ms.

TR = 5000 ms.

$f_x$: Volume fraction for x tissue of interest (WM, GM, CSF) within MRS voxel.

$conc_{H2O}$: Voxel-specific tissue water concentrations. Calculated using Equation 2.

**Equation 2.**

$$\text{conc}_{H2O} = f_{WM} \cdot [H2O]_{WM} + f_{GM} \cdot [H2O]_{GM} + f_{CSF} \cdot [H2O]_{CSF}$$

**Supplementary Tables.**

**Supplementary Table S1.** Description of functional tests.

| **Test** | **Domain assessed** | **Description** |
|---|---|---|
| Symbol digit modalities test | Processing speed | This is an iPad-based test based on the Symbol-Digit Modalities Test. Numbers from 1-9 are matched to symbols via a digit-symbol key. The participant must insert the number that matches the symbol (2 minutes). |
| 9-hole peg test | Manual dexterity | The participant must place all the pegs into the pegboard and remove them (one at a time). Two trials are performed with the dominant hand, and two with the non-dominant hand. Results are averaged for each hand, and overall. |
| Timed 25-foot walk test | Gait/mobility | Walk 25 feet as quickly (and safely) as possible. The task is repeated in the opposite direction, and the two times are averaged. |

**Supplementary Table S2.** $T_2$ relaxation times of metabolites from Swanberg et al. (2018).

*Abbreviations:* Gln, glutamine; Glu, glutamate; GSH, glutathione; mI, *myo*-inositol; ms, millisecond; NAA, *N*-acetylaspartate; NAAG, *N*-acetylaspartyl glutamate; tCho, total choline; tCr, total creatine.

| **Metabolite** | **$T_2$ (ms)** |
|---|---|
| GSH | 145 |
| NAA | 222.1 |
| NAAG | 222.1 |
| Glu | 164.1 |
| Gln | 130 |
| mI | 140.5 |
| tCho | 176.14 |
| tCr | 153 |

**Supplementary Table S3**. Ranges of GM, WM, and CSF fraction within PCC and CSWM voxels.

*Abbreviations:* CSF, cerebrospinal fluid; CSWM, centrum semiovale white matter; GM, grey matter; PCC, posterior cingulate cortex; WM, white matter.

| **Region of interest** | **GM fraction** | **WM fraction** | **CSF fraction** |
|---|---|---|---|
| PCC | 46.67-69.15% | 20.01-36.73% | 6.01-24.98% |
| CSWM | 0.77-16.82% | 69.52-99.23% | 0-1.75% |

**Supplementary Table S4.** Average CRLB of included metabolites per region of interest.

*Abbreviations:* CRLB, Cramér-rao lower bound; CSWM, centrum semiovale white matter; Gln, glutamine; Glu, glutamate; GSH, glutathione; mI, *myo*-inositol; NAA, *N*-acetylaspartate; NAAG, *N*-acetylaspartyl glutamate; PCC, posterior cingulate cortex; tCho, total choline; tCr, total creatine.

| **Metabolite** | **CRLB (%)** | |
|---|---|---|
| | **PCC** | **CSWM** |
| GSH | 16 | 12 |
| NAA | 2 | 2 |
| NAAG | 8 | 7 |
| Glu | 2 | 3 |
| Gln | 10 | 19 |
| mI | 3 | 3 |
| tCho | 5 | 3 |
| tCr | 2 | 2 |
